# WNT and TGF-Beta Pathway Alterations in Early-Onset Colorectal Cancer Among Hispanic/Latino Populations

**DOI:** 10.1101/2024.10.17.24315250

**Authors:** Cecilia Monge, Brigette Waldrup, Francisco Carranza, Enrique Velazquez-Villarreal

## Abstract

One of the fastest-growing minority groups in the U.S. is the Hispanic/Latino population. Recent studies have shown how this population is being disproportionately affected by early-onset colorectal cancer (CRC). Compared to corresponding non-Hispanic White (NHW) patients, Hispanic/Latino patients have both higher incidence of disease and rates of mortality. Two well-established drivers of early-onset CRC in the general population are alterations in the WNT and TGF-Beta signaling pathways; however, the specific roles of these pathways in Hispanic/Latino are poorly understood. Here, we assessed CRC mutations in the WNT and TGF-Beta pathways by conducting a bioinformatics analysis using cBioPortal. Cases of CRC were stratified both by age and ethnicity: 1) early-onset was defined as <50 years vs. late-onset as ≥50 years; 2) we compared early-onset in Hispanics/Latinos to early-onset in NHWs. No significant differences were evident when we compared early-onset and late-onset CRC cases within the Hispanic/Latino cohort. These results are consistent with findings from large cohorts that do not specify ethnicity. However, we found significant differences when we compared early-onset CRC in Hispanic/Latino patients to early-onset CRC in NHW patients: specifically, alterations in the gene bone morphogenetic protein-7 (BMP7) were more frequent in early-onset CRC for the Hispanic/Latino patients. In addition to these findings, we observed that both NHW patients and Hispanic/Latino patients with early-onset disease had better clinical outcomes when there was evidence of WNT pathway alterations. Conversely, only the early-onset Hispanic/Latino population had improved outcomes when there was no evidence of TGF-Beta pathway alterations. *In toto*, these findings underscore how the WNT and TGF-Beta pathways may act differently in different ethnic groups who have early-onset CRC. These findings may set a stage for developing new therapies tailored for reducing cancer health disparities.

## Introduction

Colorectal cancer (CRC) is the third most prevalent cancer in the U.S. and the second leading cause of cancer-related death globally (Sung et al., 2020). Although CRC has been historically associated with older age, recent data highlight a concerning rise in the incidence of CRC in younger individuals (<50 years) (Sinicrope, 2019) – i.e., early-onset CRC (EOCRC). For example, individuals aged 18-40 and 40-50 are increasingly being diagnosed with rectal cancer (Tait, 2023). Accordingly, other studies have shown that the incidence of EOCRC is rising at an annual rate of approximately 1.4% in individuals under 50 (Ullah, 2023). The rise in EOCRC has been observed both in the U.S. as well as globally (Ugai et al. 2022). However, the trend in early cases is especially pronounced in high-income countries like the U.S.: EOCRC now represents approximately 10% of all new CRC cases (Bhandari et al., 2017; Waddell et al., 2023). In response, the American Cancer Society recently updated its screening guidelines and lowered the recommended starting age from 50 to 45 (Ullah, 2023). To compound the urgency of detecting EOCRC, the cancer itself is associated with more advanced stages at time of diagnosis and poorer histological differentiation. Both factors contribute to worse outcomes for younger patients (Mauri et al., 2019; Siegel et al., 2020). These findings stress the importance of revising screening strategies towards early detection.

In addition to its alarming rise in the general population, EOCRC disproportionately affects one of the fastest growing minority groups in the U.S. -- the Hispanic/Latino population (Carethers et al., 2021; Shah and Camargo, 2024). This population often faces disparities in cancer outcomes and healthcare access (Goel & Boland, 2012; Monge et al., 2022). The exact cause of the disparity in EOCRC outcomes is not only poorly understood but also involves a complex interplay of multiple factors. For instance, the role of unhealthy dietary patterns, such as the high consumption of processed foods, can accelerate the development of precancerous colorectal polyps (Feng, 2023). In addition, genetic predispositions are significant contributors. Ionescu (2023) points out that EOCRC (vs. late-onset) often presents with distinct oncogenic mutations and histopathological characteristics. Importantly, only a handful of studies have comprehensively characterized the Hispanic/Latino population in terms of the molecular drivers of CRC. This population is notably underrepresented in publicly available genomic databases (Monge et al., 2024).

Two signaling pathways are well known for harboring molecular targets implicated in CRC: the WNT signaling pathway and the TGF-beta signaling pathway. While both pathways are recognized drivers of CRC progression, the molecular characteristics of early-onset CRC in Hispanic/Latino populations have not been defined sufficiently. Prior research has identified unique molecular traits in young-onset CRC: CIMP-low and LINE-1 hypomethylation (Done & Fang, 2021). Yet only a handful of reports have documented pathway-specific alterations in Hispanic/Latino patients. In this study, we aim to define the prevalence of WNT and TGF-beta pathway alterations in EOCRC among Hispanic/Latino patients, comparing these findings with both late-onset Hispanic/Latino patients and early-onset NHW patients. By leveraging bioinformatics analyses of publicly available CRC datasets, we seek to identify ethnic-specific molecular alterations in these key pathways, shed light on potential drivers of CRC in Hispanic/Latino patients, and pave the way for developing new treatment strategies for early-onset CRC in underrepresented populations.

## Methods

We assessed individual patient-level clinical and genomic data from 19 colorectal cancer (CRC) datasets available in the cBioPortal database. We included studies categorized as colorectal adenocarcinoma, colon adenocarcinoma, and rectal adenocarcinoma, along with the GENIE BPC CRC v2.0-public dataset. We excluded two studies that focused on metastatic colorectal cancer samples. After selecting the relevant studies, we filtered the samples using the following five criteria. 1) Ethnicities were filtered to include Hispanic or Latino, Spanish, NOS; Hispanic, NOS; Latino, NOS; Mexican or Spanish surname only. 2) Sample we assessed were primary tumors. 3) Cancer type was filtered to include colon adenocarcinoma, rectal adenocarcinoma, and colorectal adenocarcinoma. 4) Histology was adenocarcinoma, NOS. 5) We included only one sample per patient. This filtering process yielded three datasets that met all criteria: TCGA PanCancer Atlas, MSK Nat Commun 2022, and GENIE BPC CRC. They comprised a total of 20 early-onset and 13 late-onset CRC samples from Hispanic/Latino patients. Additionally, data regarding age at diagnosis were retrieved from each patient’s individual clinical records within the GENIE database. Alterations in the WNT and TGF-beta pathways were defined as in a previous study (Ferrell et al., 2024).

Cohorts were defined based on age categories: Early-onset patients were less than 50 years old and Late-onset patients were 50 years or older. Ethnicity was considered by classifying participants into two cohorts: Hispanics/Latinos and Non-Hispanic Whites (NHW). Within these broader categories, participants were further stratified based on the presence or absence of WNT and TGF-beta pathway alterations. This allowed us to perform a more granular analysis of the impact of these genetic factors on the cohorts. To assess differences between these defined cohorts, Chi-square tests were employed to evaluate the independence of categorical variables. This statistical approach allowed us to examine the association between age, ethnicity, and the presence of pathway alterations. Additionally, further sub-stratification was conducted based on tumor location; we distinguished colon from rectal cancers. This detailed stratification allowed us to comprehensively analyze how age, ethnicity, and tumor location interact with molecular alterations. It thus provided insights into the potential heterogeneity in patient outcomes and treatment responses within the study population. Kaplan-Meier survival curves were utilized to evaluate overall survival by assessing the influence of WNT and TGF-beta pathway alterations. This method involved estimating the survival function from time-to-event data, allowing for the visualization of survival probabilities over time. Specifically, patients were stratified based on the presence or absence of WNT and TGF-beta alterations. The resulting survival curves were plotted, and the log-rank test was employed to compare differences between cohorts, determining statistical significance. Additionally, median survival times were calculated, and 95% confidence intervals were provided to quantify the uncertainty around these estimates. This comprehensive approach enabled a clear understanding of how these molecular alterations impact patient outcomes.

## Results

The Hispanic/Latino cohort comprised 33 samples from three studies; while 36% of these patients were diagnosed at age 50 or older, 64% of them presented with EOCRC (Table 1). Colon adenocarcinoma accounted for 72% of cases, rectal adenocarcinoma for 21.2%, and colorectal adenocarcinoma for 6.1%. The cohort was predominantly male (60.6%). All samples analyzed were primary tumors to focus on the initial presentation of the cancer. At the time of diagnosis, 15.2% of patients were Stage II, 30.3% Stage III, and 39.4% Stage IV. 69.7% of patients identified as Mexican (including Chicano); 15.2% were categorized as Hispanic or Latino; 9.1% were identified by a Spanish surname only; and 6.1% classified as Spanish NOS, Hispanic NOS, or Latino NOS.

**Table 1.** Patient Demographics and Clinical Characteristics of the Hispanic/Latino (H/L) and Non-Hispanic White (NHW) cohorts.

| Clinical Feature | H/L Cohort<br>n (%) | NHW Cohort<br>n (%) |
| --- | --- | --- |
| **Age\*** |  |  |
| < 50 | 21 (63.6%) | 65 (19.0%) |
| ≥ 50 | 12 (36.4%) | 276 (80.7%) |
| **Cancer Type** |  |  |
| Colon Adenocarcinoma | 24 (72.7%) | 235 (68.7%) |
| Rectal Adenocarcinoma | 7 (21.2%) | 99 (28.9%) |
| Colorectal Adenocarcinoma | 2 (6.1%) | 8 (2.3%) |
| **Sex** |  |  |
| Male | 20 (60.6%) | 183 (53.5%) |
| Female | 13 (39.4%) | 159 (46.5%) |
| **Sample Type** |  |  |
| Primary Tumor | 33 (100.0%) | 342 (100.0%) |
| **Stage at Diagnosis \*\*** |  |  |
| Stage 0 (CIS) | 0 (0.0%) | 2 (0.6%) |
| Stage I | 0 (0.0%) | 7 (2.0%) |
| Stage II | 5 (15.2%) | 6 (1.8%) |
| Stage III | 10 (30.3%) | 22 (6.4%) |
| Stage IV | 13 (39.4%) | 14 (4.1%) |
| **Ethnicity** |  |  |
| Hispanic Or Latino | 5 (15.2%) | 0 (0.0%) |
| Mexican (includes Chicano) | 23 (69.7%) | 0 (0.0%) |
| Spanish NOS; Hispanic NOS, Latino NOS | 2 (6.1%) | 0 (0.0%) |
| Spanish surname only | 3 (9.1%) | 0 (0.0%) |
| Non-Spanish; Non-Hispanic | 0 (0.0%) | 342 (100.0%) |

\*NHW NA:1
\*\* HL NA: 5, NHW NA: 291

In comparison, the NHW cohort consisted of 342 samples from three studies, primarily comprising late-onset patients; 80.7% were aged 50 and older (Table 2). Colon adenocarcinoma was again the most common diagnosis, representing 68.7% of cases, followed by rectal adenocarcinoma (28.9%) and colorectal adenocarcinoma (2.3%). Gender distribution in this cohort showed 53.5% males. All samples were primary tumors, emphasizing initial disease presentation. The stage at diagnosis revealed that 0.6% were Stage 0 (CIS), 2% Stage I, 1.8% Stage II, 6.4% Stage III, and 4.1% Stage IV. This comprehensive demographic analysis highlights significant differences in age, cancer type, and ethnic background between the early-onset Hispanic/Latino population and the NHW cohort, underscoring important trends in colorectal cancer presentation across these populations.

**Table 2.** Age-related variations in clinical features in the Hispanic/Latino cohort. Ethnicity-associated diKerences in clinical features between early-onset Hispanic and Latino and Non-Hispanic White (NHW) cohorts.

| Clinical Feature | Early-Onset H/L<br>n (%) | Late-Onset H/L<br>n (%) | p-value | Early-Onset H/L<br>n (%) | Early-Onset NHW<br>n (%) | p-value |
| --- | --- | --- | --- | --- | --- | --- |
| Median Age (IQR) | 41 (36-45) | 63 (58-74) | 2.56E-06 | 41 (36-45) | 43 (38-47) | 0.2149 |
| Median mutation count | 9 (6-10) | 24 (7-63) | 0.4414 | 9 (6-10) | 58 (7-79) | 0.01093 |
| Median TMB (IQR) * | 4.87 (1.89-21.38) | 3.8 (3.22-4.82) | 1 | 4.87 (1.89-21.38) | 4.32 (2.53-7.56) | 0.9215 |
| Median FGA** | 0.06 (0.04-0.28) | 0.31 (0.10-0.36) | 0.184 | 0.06 (0.04-0.28) | 0.17 (0.06-0.34) | 0.2275 |
| Oncotree Code |  |  |  |  |  |  |
| COAD | 14 (66.7%) | 10 (83.3%) | 0.56 | 14 (66.7%) | 38 (55.1%) | 0.22336 |
| COADREAD | 2 (9.5%) | 0 (0.0%) |  | 2 (9.5%) | 5 (7.2%) |  |
| READ | 5 (23.8%) | 2 (16.7%) |  | 5 (23.8%) | 22 (31.9%) |  |
| Sex |  |  |  |  |  |  |
| Male | 12 (57.1%) | 8 (66.7%) | 0.7188 | 12 (57.1%) | 35 (50.7%) | 0.9906 |
| Female | 9 (42.9%) | 4 (33.3%) |  | 9 (42.9%) | 30 (43.5%) |  |
| APC Mutation |  |  |  |  |  |  |
| Present | 14 (66.7%) | 11 (91.7%) | 0.2062 | 14 (66.7%) | 36 (52.2%) | 0.5114 |
| Absent | 7 (33.3%) | 1 (8.3%) |  | 7 (33.3%) | 29 (42.0%) |  |
| KRAS Mutation |  |  |  |  |  |  |
| Present | 8 (38.1%) | 6 (50.0%) | 0.7157 | 8 (38.1%) | 26 (37.7%) | 1 |
| Absent | 13 (61.9%) | 6 (50.0%) |  | 13 (61.9%) | 39 (56.5%) |  |
| TP53 Mutation |  |  |  |  |  |  |
| Present | 15 (71.4%) | 9 (75.0%) | 1 | 15 (71.4%) | 47 (68.1%) | 1 |
| Absent | 6 (28.6%) | 3 (25.0%) |  | 6 (28.6%) | 18 (26.1%) |  |
| BMP7 Mutation |  |  |  |  |  |  |
| Present | 0 (0.0%) | 0 (0.0%) | 1 | 0 (0.0%) | 12 (18.5%) | 0.03361 |
| Absent | 21 (100.0%) | 12 (100.0%) |  | 21 (100.0%) | 53 (81.5%) |  |
| MTOR Mutation |  |  |  |  |  |  |
| Present | 3 (14.3%) | 1 (8.3%) | 1 | 3 (14.3%) | 1 (1.5%) | 0.04353 |
| Absent | 18 (85.7%) | 11 (91.7%) |  | 18 (85.7%) | 64 (98.5%) |  |

\*EO H/L NA: 17, LO H/L NA: 9, EO NHW NA: 42
\*\*EO H/L NA: 4, EO NHW NA: 1

In our Hispanic/Latino cohort when comparing early-onset with late onset, the patients with EOCRC were similarly predominantly male compared to patients over 50 years of age (57.1 vs. 66.7%, p=1). The patients with EOCRC were similarly frequently female compared to patients over 50 years of age (42.8% vs. 33.3%, p=0.9). The median tumor mutational burden (TMB) was significantly higher in patients with EOCRC (median: 4.87, IQR: 1.89–21.38) compared to patients > 50 years of age (median: 3.8, IQR: 3.22-4.82) (p = 1). Patients with young-onset CRC (≤ 50 years old) had lower rates of TP53 (71.4% versus 75%, *p* = 1), APC (66.7 versus 91.7%, p = 0.2), and KRAS (38.1% versus 50%, p < 0.7) compared to patients over 50 years of age, however this was not statistically significant (Table 3).

**Table 3.**
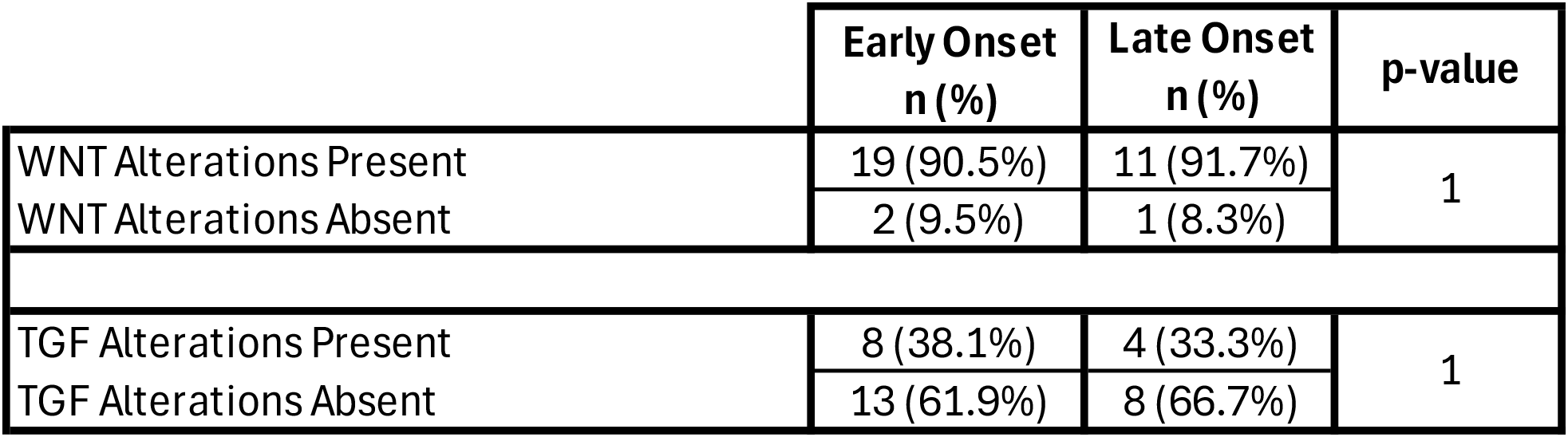
Rates of WNT and TGF-Beta pathway alterations among early-onset and late-onset Hispanic/Latino CRC patients.

In our early-onset Hispanic/Latino cohort when comparing with Non-Hispanic White Cohort, the Hispanic/Latino patients with EOCRC were less frequently male compared to early-onset NHW (57.1 vs. 50.7%, p=1). The patients with early-onset CRC were similarly frequently female compared to early-onset NHW patients (42.9% vs. 43.5%, p=0.9). The median tumor mutational burden (TMB) was significantly higher in patients with early-onset Hispanic/Latino CRC patients (median: 4.87, IQR: 1.89–21.38) compared to early-onset NHW patients (median: 4.32, IQR: 2.53-7.56) (p = 1). Hispanics/Latinos patients with early-onset CRC had higher rates of TP53 (71.4% versus 68.1%, *p* = 1), APC (66.7 versus 52.2%, p = 0.5), and KRAS (38.1% versus 37.7%, p < 0.7) compared to early onset NHW patients, however this was not statistically significant (Table 4).

**Table 4.** Rates of WNT and TGF-Beta pathway alterations among early-onset Hispanic/Latino and Non-Hispanic White (NHW) CRC Patients.

|  | Early Onset H/L<br>n (%) | Early Onset NHW<br>n (%) | p-value |
| --- | --- | --- | --- |
| WNT Alterations Present | 19 (90.5%) | 44 (67.7%) | 0.04876 |
| WNT Alterations Absent | 2 (9.5%) | 21 (32.3%) |  |
| TGF Alterations Present | 8 (38.1%) | 17 (26.2%) | 0.4071 |
| TGF Alterations Absent | 13 (61.9%) | 48 (73.8%) |  |

The Kaplan-Meier survival analysis for Hispanic/Latino EOCRC patients showed no significant difference in overall survival between those with and without WNT pathway alterations (Figure 2). Both groups exhibited a similar decline in survival probability over time, with overlapping confidence intervals. Although patients with WNT pathway alterations experienced a more gradual decline in survival probability compared to those without alterations, who had a sharper decrease within the first 10 months, the survival curves converged by the end of the observation period. The p-value of 0.72 indicated that the difference in survival outcomes was not statistically significant, suggesting that WNT pathway alterations do not substantially impact overall survival in this cohort of early-onset Hispanic/Latino CRC patients. Similarly, Kaplan-Meier survival analysis comparing overall survival in early-onset Hispanic/Latino CRC patients with and without TGF-beta pathway alterations revealed no significant difference between the groups (Figure 2). The survival curves followed a nearly identical trajectory, with overlapping confidence intervals that indicated comparable survival probabilities over time.

**Figure 1.**
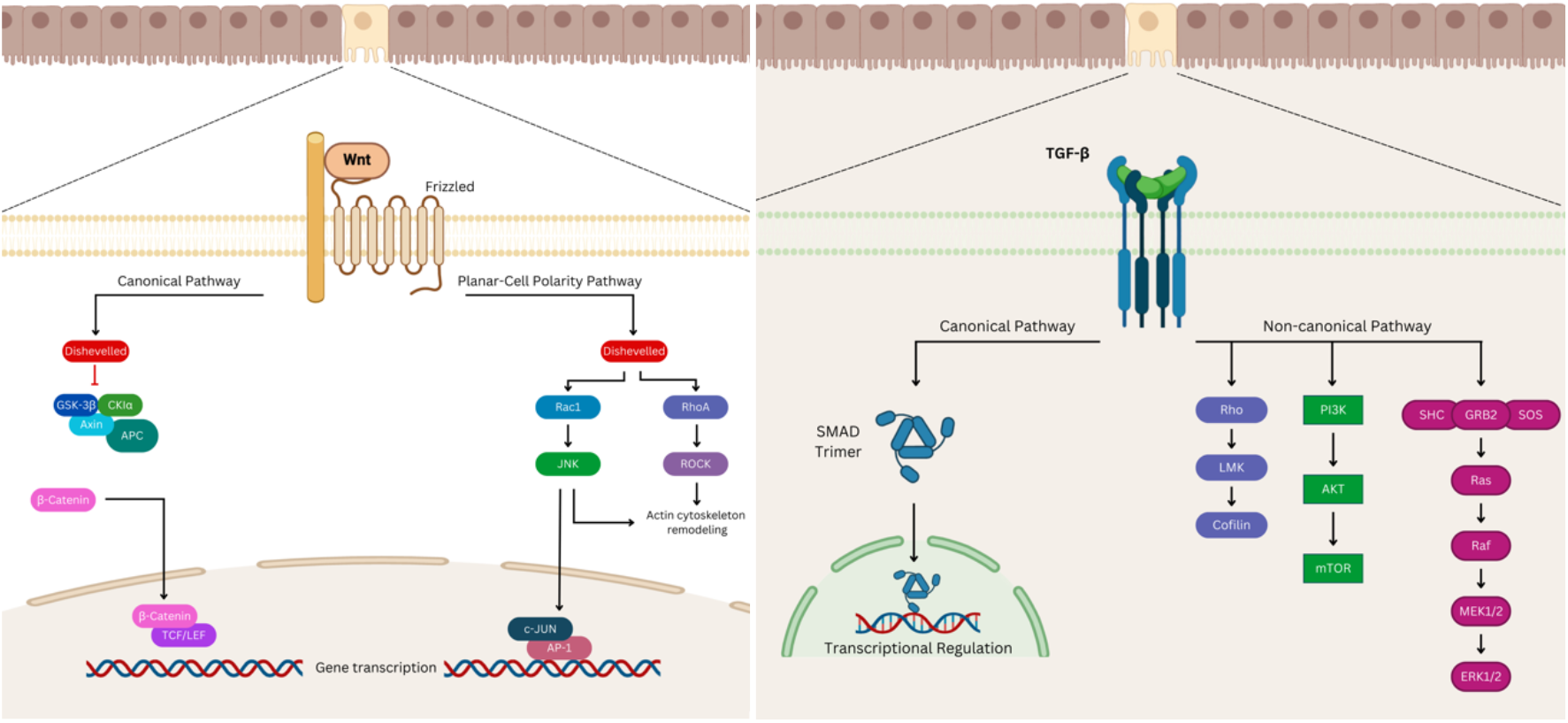
Illustration of the WNT signaling pathway on the left and the TGF-beta signaling pathway on the right.

**Figure 2.**
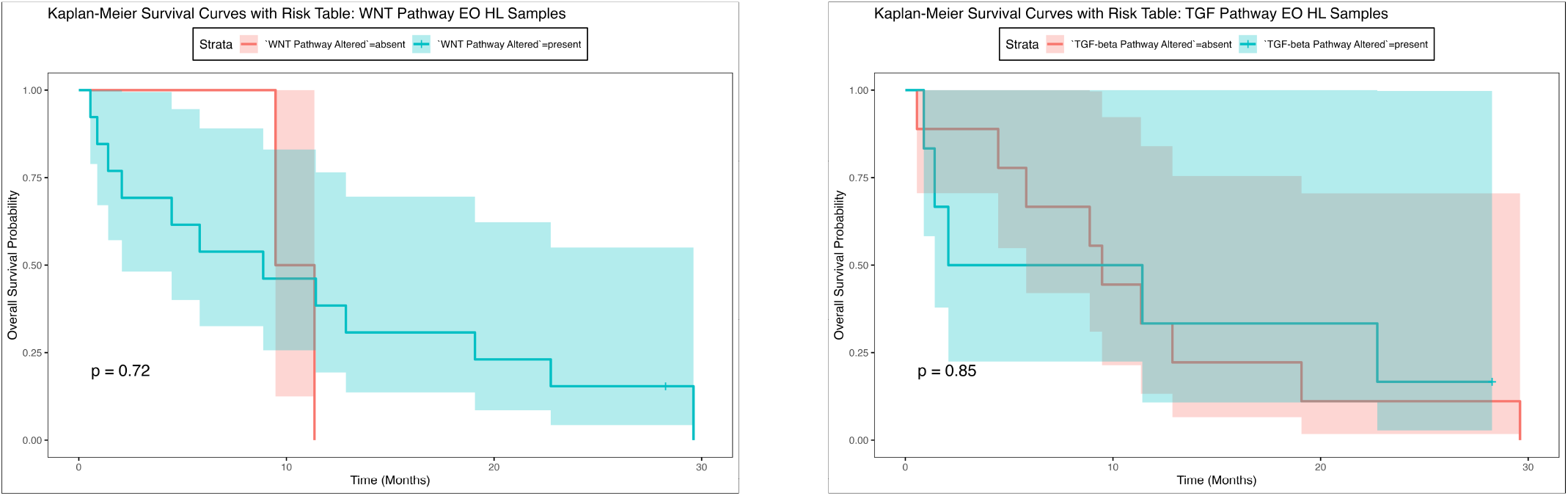
Overall survival curves of early-onset Hispanic/Latino patients stratified by the presence or absence of WNT (left) and TGF-β (right) pathway alterations.

Patients with TGF-beta pathway alterations showed a slightly slower decline in survival probability early in the observation period, but the curves converged by the end. The p-value of 0.85 confirmed that the difference in survival between patients with and without TGF-beta pathway alterations was not statistically significant these results suggest that alterations in the TGF-beta pathway do not significantly influence overall survival in this cohort of early-onset Hispanic/Latino CRC patients. Similar to NHW overall survival results (Supplementary Figure 1), these findings highlight that, in this specific ethnic cohort, neither WNT nor TGF-beta pathway alterations are major determinants of survival outcomes in early-onset colorectal cancer.

In our Hispanic/Latino cohort, there was no significant difference in the frequency of WNT pathway alterations between early-onset and late-onset colorectal cancer (CRC) patients (90.5% versus 91.7%, p = 1) (Table 5). However, the rate of APC alterations was lower in early-onset CRC compared to late-onset patients, though this difference was not statistically significant (66.7% versus 91.7%, p = 0.2) (Supplementary Table 1). Notably, several WNT pathway-related genes, including AXIN1, AXIN2, and RNF43, exhibited alterations in early-onset patients but no alterations in late-onset CRC patients. GSK3B was the only WNT pathway-related gene that showed no alterations in either age group. When stratified by cancer type (e.g., colon versus rectum adenocarcinoma), no significant differences in WNT pathway alterations were observed, indicating that these genetic variations are consistent across CRC subtypes within this ethnic cohort (Supplementary Table 2). Similarly, TGF-Beta pathway alterations were not significant between early-onset and late-onset patients in the Hispanic/Latino cohort, nor did stratification by cancer type reveal any substantial variation in these pathway alterations. This suggests that, while some WNT pathway genes are more commonly altered in early-onset cases, these differences are largely independent of the specific CRC subtype.

When comparing early-onset Hispanic/Latino patients with early-onset NHW patients, significant differences in WNT pathway alterations were observed (**90.5% vs 67.7%, p < 0.05**) (Table 6). The frequency of APC alterations was lower in early-onset Hispanic/Latino patients compared to NHW patients, though this difference was not statistically significant (66.7% vs 55.4%, p = 0.5) (Supplementary Table 3). Alterations in AXIN1 and AXIN2 were similar between the two ethnic groups, while mutations in RNF43 were more frequent in early-onset Hispanic/Latino patients than in their NHW counterparts (23.8% vs 7.7%, p = 0.1). When stratified by cancer type, early-onset Hispanic/Latino patients with colon adenocarcinoma showed a trend toward marginal significance in WNT pathway alterations compared to early-onset NHW patients (93.3% vs 69%, p = 0.08), suggesting potential ethnic-specific variations in this pathway for colon cancer (Supplementary Table 4). Although no significant differences were observed in overall TGF-Beta pathway alterations, a notable difference in BMP7 mutations was found, with early-onset Hispanic/Latino patients showing no BMP7 alterations, compared to 18.5% of early-onset NHW patients (p < 0.03). These findings highlight key ethnic disparities in molecular pathways, particularly within the WNT and TGF-Beta pathways, associated with early-onset colorectal cancer.

## Discussion

The WNT signaling pathway is essential for regulating stem cell renewal and cell fate in the intestinal epithelium (Bugter et al., 2021; Zhang et al., 2020; Gamage et al., 2024; Peng et al., 2015). WNT signaling is frequently dysregulated in CRC due to mutations in tumor suppressor genes: three examples are APC, AXIN1, and AXIN2. Such mutations lead to sustained WNT signaling, may grant self-renewing properties to cells, and ultimately can promote tumor progression (Bugter et al., 2021; Wickström et al., 2015; Zhang et al., 2022). In studies across broad patient populations, WNT signaling was a key driver of colonic carcinogenesis (Bugter et al., 2021; Sun et al., 2024; Tang et al., 2020). Additionally, WNT signaling appears to be a promising target for reducing cancer stem cell characteristics (Wei et al., 2021). Such cells are considered an important subpopulation of cancer cells to target with new therapies because they are resistance to traditional chemotherapy and may be responsible for relapse.

The TGF-beta signaling pathway similarly plays a crucial role in regulating cell proliferation and apoptosis in normal intestinal tissues. Under normal physiological conditions, TGF-beta functions as a tumor suppressor by inducing cell cycle arrest and promoting programmed cell death (Xu et al., 2007; Bellam et al., 2010). Conversely, aberrantly elevated TGF-beta levels have been strongly linked to both progression and poor clinical outcomes in CRC (Pastini et al., 2024; Shi et al. 2023). High TGF-beta expression has been correlated with advanced disease stages, greater recurrence risk, and poorer clinical outcomes (Heldin et al., 2012; Moustakas et al., 2012; Ehata et al., 2020). Mechanistically, the progression of CRC is exacerbated by mutations in TGF-beta pathway genes like BMP7; this mutation allows tumor cells to escape growth-inhibitory effects and evade apoptosis (Zhou et al., 2023; Li et al., 2020; Liu et al., 2017; Ren et al., 2016). Taken altogether, both WNT and TGF-beta play well-documented roles in CRC for the general population of patients. Nevertheless, merely a handful of studies have probed how these pathways operate in Hispanic/Latino populations with EOCRC.

In our studies, ethnicity in EOCRC appeared to have an impact on WNT signaling. For example, our Hispanic/Latino patients (vs. NHW) had marginally higher rates of WNT pathway alterations. This was especially true in early-onset colon cancer (vs. rectal), which suggests a potential interaction between tumor site and ethnicity in CRC pathogenesis. Other, specific trends we observed in WNT pathway mutations in Hispanic/Latino patients include alterations in AXIN1, AXIN2, and RNF43. Interestingly, our study revealed no significant differences in the prevalence of APC, TP53, or KRAS mutations between early-onset Hispanic/Latino and NHW patients. These results are consistent with previous studies that suggest a lower frequency of APC mutations in younger CRC patients, particularly in early-onset CRC cases (Done & Fang, 2021). While our study provides valuable insights into the impact of ethnicity on molecular characteristics of EOCRC, it has several limitations which are important to note: the retrospective nature of the bioinformatics analysis, combined with potential selection bias from publicly available genomic databases, may limit the generalizability of our findings. Furthermore, the underrepresentation of Hispanic/Latino in these databases (Monge et al., 2022; Wilson et al 2006) hinders our ability to draw comprehensive conclusions regarding molecular disparities in CRC across all Hispanic/Latino subpopulations. Larger, prospective studies are needed to validate our findings and explore the underlying biological mechanisms driving these ethnic differences in CRC. Nevertheless, our results appear to align with emerging studies in this arena.

Recent studies have revealed other types of significant variations in molecular patterns among different ethnic groups. For instance, Pérez-Mayoral et al. (2023) identified molecular and sociodemographic disparities in Hispanic/Latino populations, particularly in Puerto Rico, where genetic ancestry was shown to be a key factor in CRC development. Their findings indicated that individuals with higher levels of African ancestry had a greater likelihood of developing rectal tumors. Similarly, Yamada et al. (2023) reported notable differences in differentially expressed genes (DEGs) and pathway utilization among White Americans, Alabama African Americans, and Oklahoma American Indians. These results suggest that distinct biological patterns in CRC may arise from ethnic and racial differences shaped by factors like diet, geography, and cultural practices. Therefore, ethnic diversity is crucial in CRC research to develop personalized strategies for prevention, diagnosis, and treatment.

In our studies, the frequency of BMP7 mutations was notably higher in early-onset Hispanic/Latino patients. Alterations in BMP7, a gene within the TGF-beta signaling pathway, were significantly more frequent in early-onset Hispanic/Latino patients (vs. NHWs). Accordingly, in prior studies, TGF-beta pathway dysregulation was associated with poorer clinical outcomes in CRC (Heldin et al., 2012; Ehata et al., 2020; Khalili-Tanha et al., 2023). Interestingly, our data also suggest that the absence of TGF-beta pathway alterations in early-onset Hispanic/Latino patients is linked to improved survival; contrastingly, NHW patients with TGF-beta alterations are more strongly associated with aggressive disease. This difference may reflect ethnic-specific tumor biology and highlight the importance of tailored therapeutic strategies for Hispanic/Latino populations. The elevated TMB observed in early-onset Hispanic/Latino patients compared to late-onset patients suggests increased genomic instability in younger individuals, which may contribute to the higher incidence and poorer prognosis in this population. High TMB has been associated with better responses to immunotherapy, and these findings may have clinical implications for the development of targeted therapies in Hispanic/Latino CRC patients (Shah & Camargo, 2024).

## Conclusion

Our study highlights significant ethnic disparities in WNT and TGF-beta pathway alterations in EOCRC between Hispanic/Latino and NHW populations. The higher prevalence of WNT pathway alterations and TGF-B-pathway related gene BMP7 mutations in early-onset Hispanic/Latino CRC patients underscores the need for further investigation into the unique molecular drivers of CRC in this underserved population. As precision medicine continues to evolve, it is essential to address these disparities to develop more effective, personalized treatment strategies that improve clinical outcomes for Hispanic/Latino CRC patients.

## Data Availability

All data produced are available online at cbioportal.org and are available upon reasonable request to the authors.

https://www.cbioportal.org/

https://genie.cbioportal.org/?continue

---

**Supplementary Figure 1.**
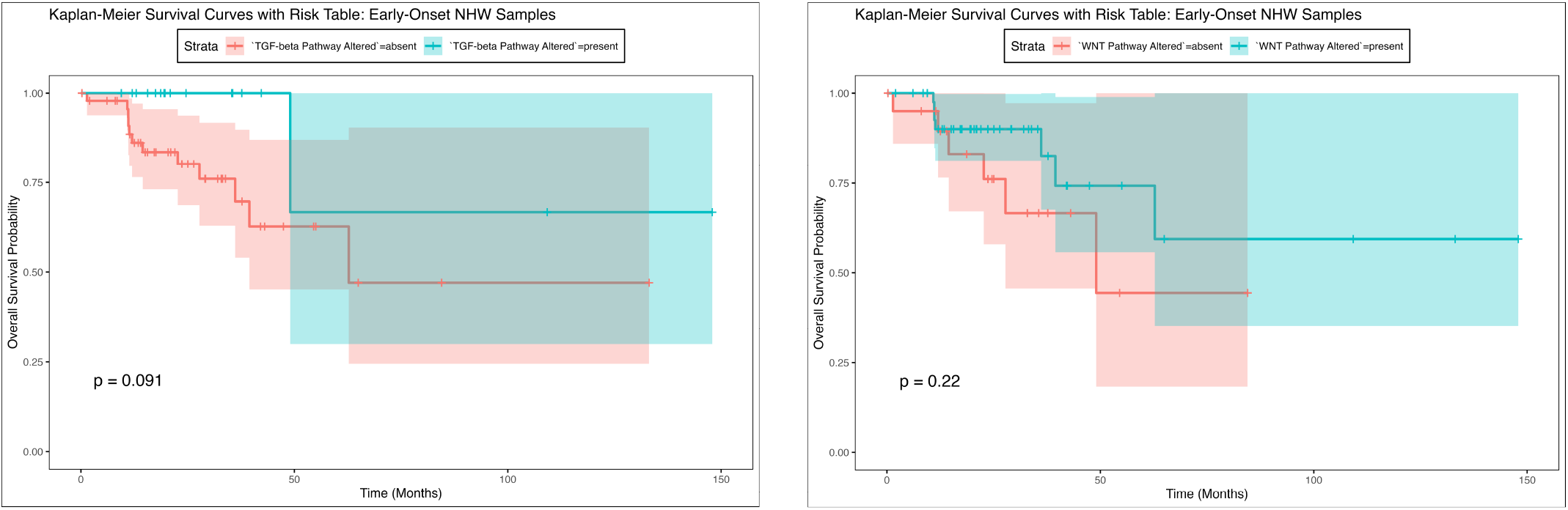
Overall survival curves of early-onset Non-Hispanic White (NHW) patients stratified by the presence or absence of WNT (left) and TGF-β (right) pathway alterations.

**Supplementary Table 1.**
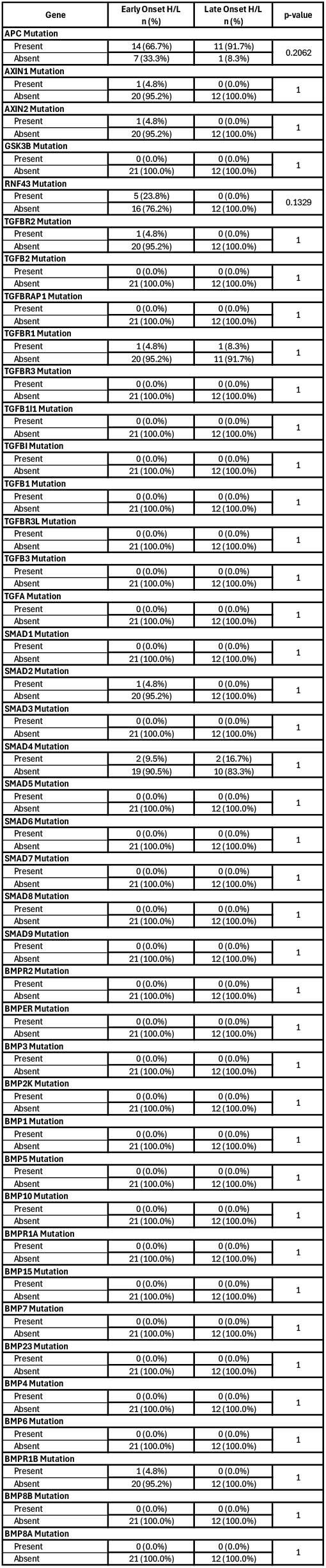
Alteration Rates of WNT and TGF-Beta Pathway-Related Genes Among Early-Onset and Late-Onset Hispanic/Latino CRC Patients.

**Supplementary Figure 2.**
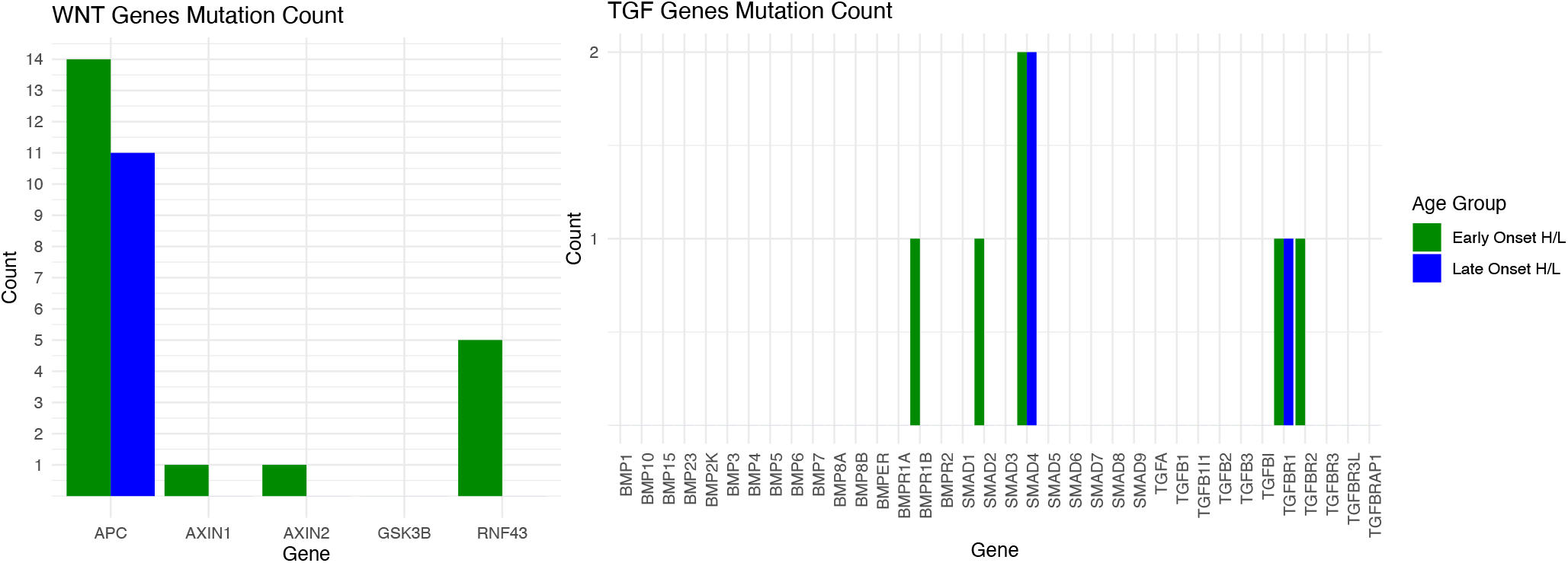
Counts of WNT and TGF-Beta Pathway-Related Gene Alterations in Early-Onset and Late-Onset Hispanic/Latino CRC Patients.

**Supplementary Table 2.**
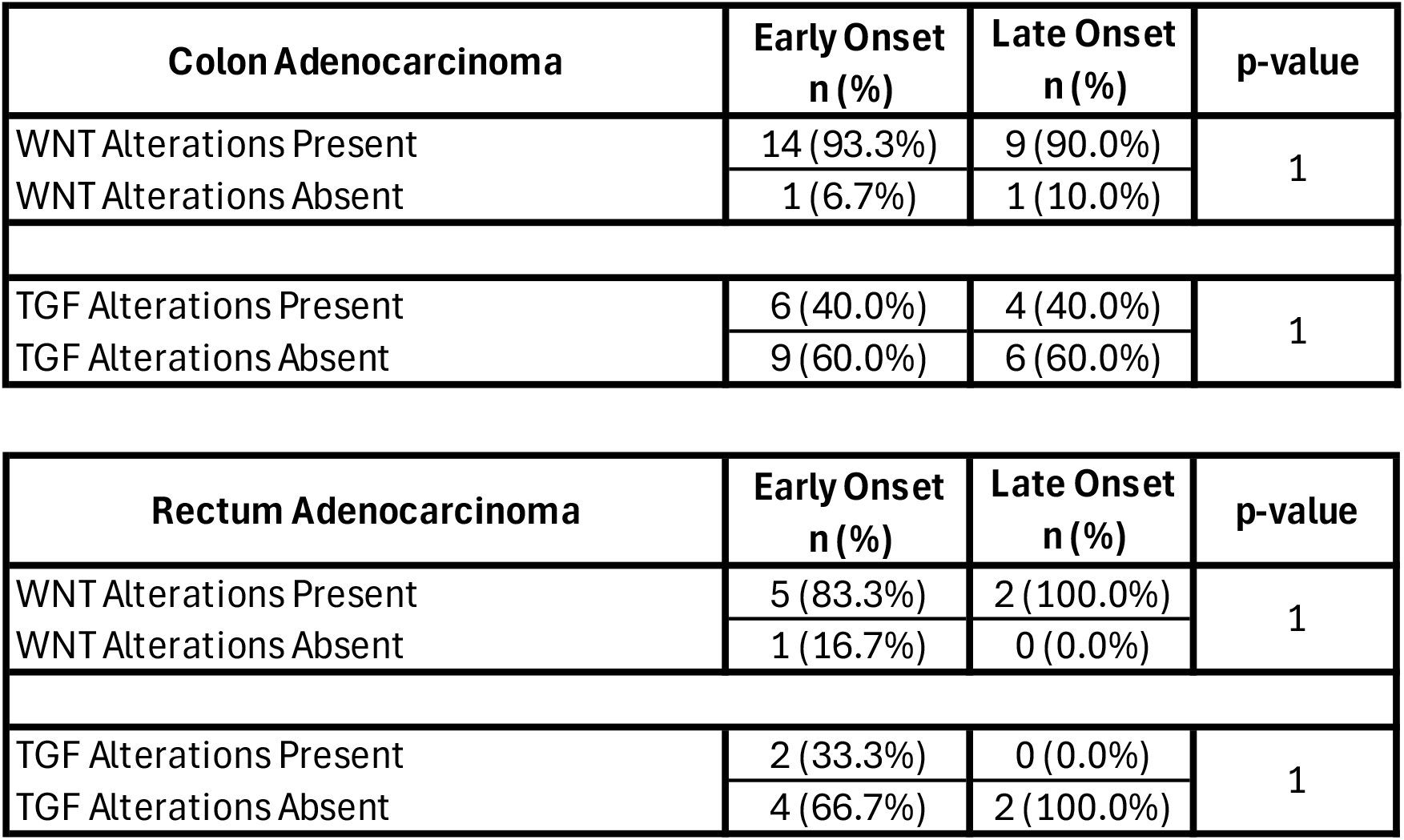
Rates of WNT and TGF-Beta pathway alterations in early-onset and late-onset Hispanic/Latino CRC patients, stratified by colon and rectal adenocarcinomas.

**Supplementary Table 3.**
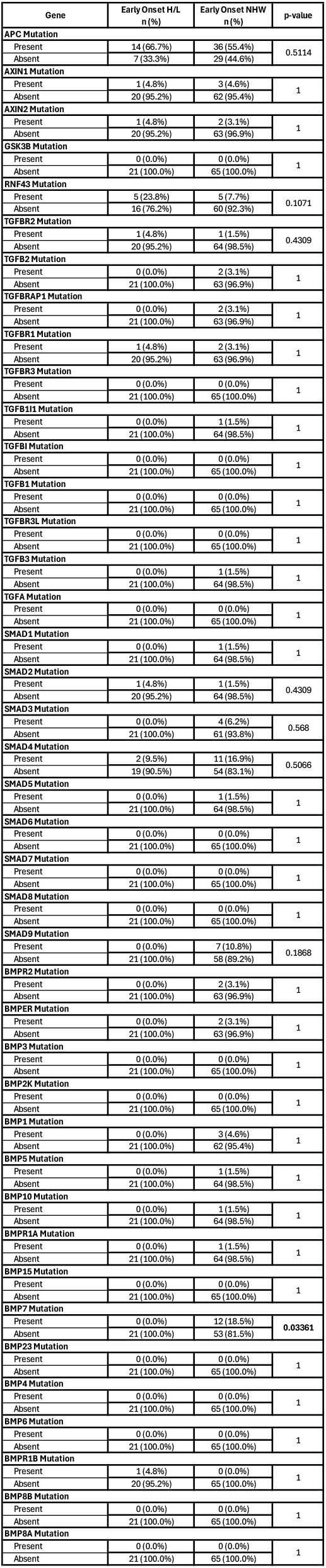
Alteration Rates of WNT and TGF-Beta Pathway-Related Genes in Early-Onset Hispanic/Latino and Non-Hispanic White CRC Patients.

**Supplementary Figure 3.**
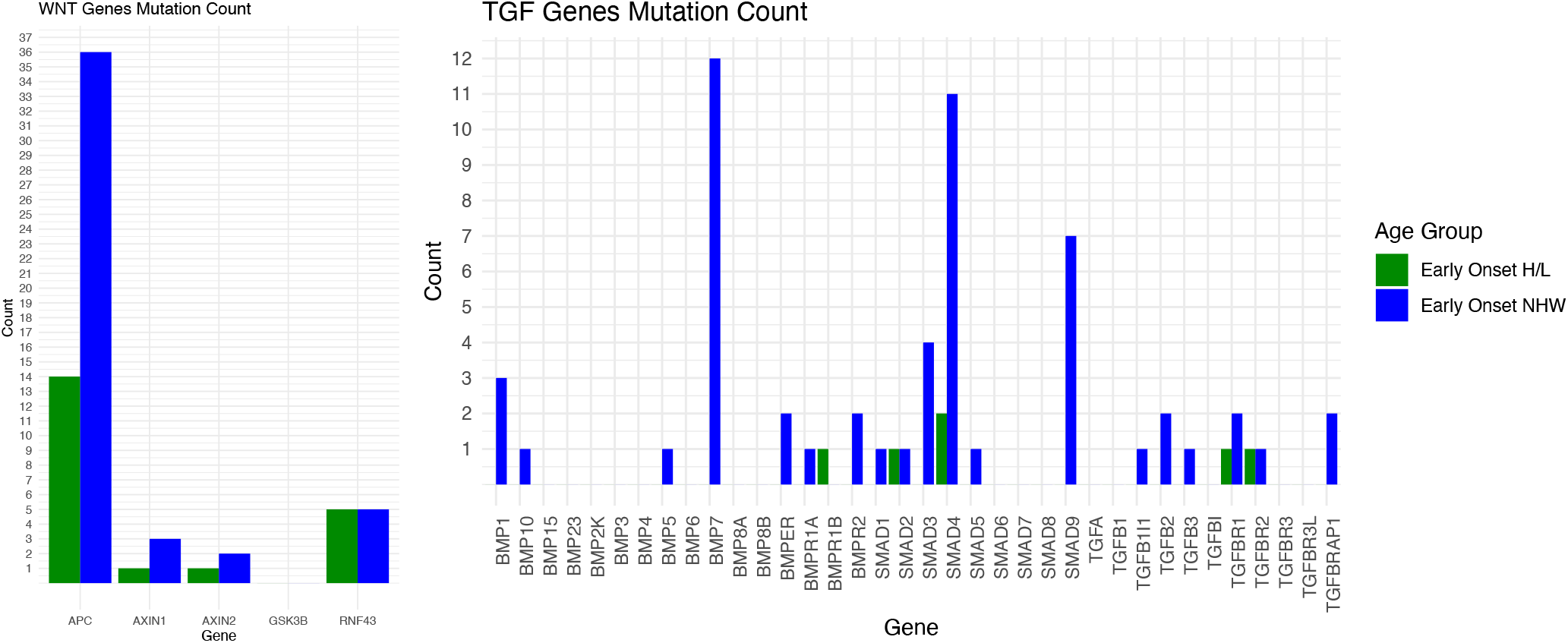
Counts of WNT and TGF-Beta Pathway-Related Gene Alterations in Early-Onset Hispanic/Latino and Non-Hispanic White CRC Patients.

**Supplementary Table 4.** Rates of WNT and TGF-Beta pathway alterations in early-onset Hispanic/Latino and Non-Hispanic White (NHW) CRC patients, stratified by colon and rectal adenocarcinomas.

| Colon Adenocarcinoma | Early-Onset H/L<br>n (%) | Early-Onset NHW<br>n (%) | p-value |
| --- | --- | --- | --- |
| WNT Alterations Present | 14 (93.3%) | 29 (69.0%) | 0.08416 |
| WNT Alterations Absent | 1 (6.7%) | 13 (31.0%) |  |
| TGF Alterations Present | 6 (40.0%) | 11 (26.2%) | 0.4998 |
| TGF Alterations Absent | 9 (60.0%) | 31 (73.8%) |  |

| Rectum Adenocarcinoma | Early-Onset H/L<br>n (%) | Early-Onset NHW<br>n (%) | p-value |
| --- | --- | --- | --- |
| WNT Alterations Present | 5 (83.3%) | 15 (65.2%) | 0.6328 |
| WNT Alterations Absent | 1 (16.7%) | 8 (34.8%) |  |
| TGF Alterations Present | 2 (33.3%) | 6 (26.1%) | 1 |
| TGF Alterations Absent | 4 (66.7%) | 17 (73.9%) |  |

## References

1. Bhandari A, Woodhouse M, Gupta S. Colorectal cancer is a leading cause of cancer incidence and mortality among adults younger than 50 years in the USA: a SEER-based analysis with comparison to other young-onset cancers. J Investig Med. 2017 Feb;65(2):311–315. doi: 10.1136/jim-2016-000229. Epub 2016 Nov 18. PMID: 27864324; PMCID: PMC5564445.

2. Bellam N, Pasche B. Tgf-beta signaling alterations and colon cancer. Cancer Treat Res. 2010;155:85–103. doi: 10.1007/978-1-4419-6033-7_5. PMID: 20517689.

3. Bugter JM, Fenderico N, Maurice MM. Mutations and mechanisms of WNT pathway tumour suppressors in cancer. Nat Rev Cancer. 2021 Jan;21(1):5–21. doi: 10.1038/s41568-020-00307-z. Epub 2020 Oct 23. Erratum in: Nat Rev Cancer. 2021 Jan;21(1):64. doi: 10.1038/s41568-020-00316-y. PMID: 33097916.

4. Carethers JM. Racial and ethnic disparities in colorectal cancer incidence and mortality. Adv Cancer Res. 2021;151:197–229. doi: 10.1016/bs.acr.2021.02.007. Epub 2021 May 5. PMID: 34148614; PMCID: PMC9069392.

5. Done JZ, Fang SH. Young-onset colorectal cancer: A review. World J Gastrointest Oncol. 2021 Aug 15;13(8):856–866. doi: 10.4251/wjgo.v13.i8.856. PMID: 34457191; PMCID: PMC8371519.

6. Ehata S, Miyazono K. Bone Morphogenetic Protein Signaling in Cancer; Some Topics in the Recent 10 Years. Front Cell Dev Biol. 2022 May 25;10:883523. doi: 10.3389/fcell.2022.883523. PMID: 35693928; PMCID: PMC9174896.

7. Feng CH, Zhang Q, Chen J, Mao LQ, Sun Q, He Y, Yao LH. Factors influencing age at onset of colorectal polyps and benefit-finding after polypectomy. Medicine (Baltimore). 2023 Sep 29;102(39):e35336. doi: 10.1097/MD.0000000000035336. PMID: 37773792; PMCID: PMC10545222.

8. Gamage CDB, Kim JH, Zhou R, Park SY, Pulat S, Varlı M, Nam SJ, Kim H. Plectalibertellenone A suppresses colorectal cancer cell motility and glucose metabolism by targeting TGF-β/Smad and Wnt pathways. Biofactors. 2024 Sep 18. doi: 10.1002/biof.2120. PMID: 39291722.

9. Goel A, Boland CR. Epigenetics of colorectal cancer. Gastroenterology. 2012 Dec;143(6):1442-1460.e1. doi: 10.1053/j.gastro.2012.09.032. Epub 2012 Sep 20. PMID: 23000599; PMCID: PMC3611241.

10. Heldin CH, Vanlandewijck M, Moustakas A. Regulation of EMT by TGFβ in cancer. FEBS Lett. 2012 Jul 4;586(14):1959–70. doi: 10.1016/j.febslet.2012.02.037. PMID: 22710176.

11. Ionescu VA, Gheorghe G, Bacalbasa N, Chiotoroiu AL, Diaconu C. Colorectal Cancer: From Risk Factors to Oncogenesis. Medicina (Kaunas). 2023 Sep 12;59(9):1646. doi: 10.3390/medicina59091646. PMID: 37763765; PMCID: PMC10537191.

12. Khalili-Tanha G, Fiuji H, Gharib M, Moghbeli M, Khalili-Tanha N, Rahmani F, Shakour N, Maftooh M, Hassanian SM, Asgharzadeh F, Shahidsales S, Anvari K, Mozafari MR, Ferns GA, Batra J, Giovannetti E, Khazaei M, Avan A. Dual targeting of TGF-β and PD-L1 inhibits tumor growth in TGF-β/PD-L1-driven colorectal carcinoma. Life Sci. 2023 Sep 1;328:121865. doi: 10.1016/j.lfs.2023.121865. PMID: 37336360.

13. Li Q, Ma Y, Liu XL, Mu L, He BC, Wu K, Sun WJ. Anti proliferative effect of honokiol on SW620 cells through upregulating BMP7 expression via the TGF β1/p53 signaling pathway. Oncol Rep. 2020 Nov;44(5):2093–2107. doi: 10.3892/or.2020.7745. PMID: 32901874; PMCID: PMC7551181.

14. Liu RX, Ren WY, Ma Y, Liao YP, Wang H, Zhu JH, Jiang HT, Wu K, He BC, Sun WJ. BMP7 mediates the anticancer effect of honokiol by upregulating p53 in HCT116 cells. Int J Oncol. 2017 Sep;51(3):907–917. doi: 10.3892/ijo.2017.4078. PMID: 28731124.

15. Mauri G, Sartore-Bianchi A, Russo AG, Marsoni S, Bardelli A, Siena S. Early-onset colorectal cancer in young individuals. Mol Oncol. 2019 Feb;13(2):109–131. doi: 10.1002/1878-0261.12417. Epub 2018 Dec 22. PMID: 30520562; PMCID: PMC6360363.

16. Monge C, Greten TF. Underrepresentation of Hispanics in clinical trials for liver cancer in the United States over the past 20 years. Cancer Med. 2024 Jan;13(1). doi: 10.1002/cam4.6814. Epub 2023 Dec 20. PMID: 38124450; PMCID: PMC10807616.

17. Monge C, Xie C, Myojin Y, Coffman K, Hrones DM, Wang S, Hernandez JM, Wood BJ, Levy EB, Juburi I, Hewitt SM, Kleiner DE, Steinberg SM, Figg WD, Redd B, Homan P, Cam M, Ruf B, Duffy AG, Greten TF. Phase I/II study of PexaVec in combination with immune checkpoint inhibition in refractory metastatic colorectal cancer. J Immunother Cancer. 2023 Feb;11(2). doi: 10.1136/jitc-2022-005640. PMID: 36754451; PMCID: PMC9923269.

18. Moustakas A, Heldin CH. Induction of epithelial-mesenchymal transition by transforming growth factor β. Semin Cancer Biol. 2012 Oct;22(5-6):446–54. doi: 10.1016/j.semcancer.2012.04.002. PMID: 22548724.

19. Pastini, A. (2024). Role of the tgf-beta/smad pathway in tumor radioresistance to boron neutron capture therapy (bnct) in a human colon carcinoma cell line. 10.21203/rs.3.rs-4497846/v1

20. Peng X, Luo Z, Kang Q, Deng D, Wang Q, Peng H, Wang S, Wei Z. FOXQ1 mediates the crosstalk between TGF-β and Wnt signaling pathways in the progression of colorectal cancer. Cancer Biol Ther. 2015;16(7):1099–109. doi: 10.1080/15384047.2015.1047568. PMID: 25955104; PMCID: PMC4623466.

21. Perez-Mayoral J, Gonzalez-Pons M, Centeno-Girona H, Montes-Rodríguez IM, Soto-Salgado M, Suárez B, Rodríguez N, Colón G, Sevilla J, Jorge D, Llor X, Xicola RM, Toro DH, Tous-López L, Torres-Torres M, Reyes JS, López-Acevedo N, Goel A, Rodríguez-Quilichini S, Cruz-Correa M. Molecular and Sociodemographic Colorectal Cancer Disparities in Latinos Living in Puerto Rico. Genes (Basel). 2023 Apr 11;14(4):894. doi: 10.3390/genes14040894. PMID: 37107652; PMCID: PMC10138302.

22. Ren CM, Li Y, Chen QZ, Zeng YH, Shao Y, Wu QX, Yuan SX, Yang JQ, Yu Y, Wu K, He BC, Sun WJ. Oridonin inhibits the proliferation of human colon cancer cells by upregulating BMP7 to activate p38 MAPK. Oncol Rep. 2016 May;35(5):2691–8. doi: 10.3892/or.2016.4654. PMID: 26986967.

23. Shah SC, Camargo MC, Lamm M, Bustamante R, Roumie CL, Wilson O, Halvorson AE, Greevy R, Liu L, Gupta S, Demb J. Impact of Helicobacter pylori Infection and Treatment on Colorectal Cancer in a Large, Nationwide Cohort. J Clin Oncol. 2024 Jun 1;42(16):1881–1889. doi: 10.1200/JCO.23.00703. Epub 2024 Mar 1. PMID: 38427927.

24. Shi, L., Wang, Y., Cheng, Z., Lv, Z., Lu, R., & Gao, H. (2023). The synergistic effect of tgm2 and tgfβ2 on the prognosis of colon cancer patients. 10.21203/rs.3.rs-2781254/v1

25. Siegel RL, Miller KD, Goding Sauer A, Fedewa SA, Butterly LF, Anderson JC, Cercek A, Smith RA, Jemal A. Colorectal cancer statistics, 2020. CA Cancer J Clin. 2020 May;70(3):145–164. doi: 10.3322/caac.21601. Epub 2020 Mar 5. PMID: 32133645.

26. Sinicrope FA. Increasing Incidence of Early-Onset Colorectal Cancer. N Engl J Med. 2022 Apr 21;386(16):1547–1558. doi: 10.1056/NEJMra2200869. PMID: 35443109.

27. Sun L, Xing J, Zhou X, Song X, Gao S. Wnt/β-catenin signalling, epithelial-mesenchymal transition and crosslink signalling in colorectal cancer cells. Biomed Pharmacother. 2024 Jun;175:116685. doi: 10.1016/j.biopha.2024.116685. PMID: 38710151.

28. Sung H, Ferlay J, Siegel RL, Laversanne M, Soerjomataram I, Jemal A, Bray F. Global Cancer Statistics 2020: GLOBOCAN Estimates of Incidence and Mortality Worldwide for 36 Cancers in 185 Countries. CA Cancer J Clin. 2021 May;71(3):209–249. doi: 10.3322/caac.21660. Epub 2021 Feb 4. PMID: 33538338.

29. Tang Q, Chen J, Di Z, Yuan W, Zhou Z, Liu Z, Han S, Liu Y, Ying G, Shu X, Di M. TM4SF1 promotes EMT and cancer stemness via the Wnt/β-catenin/SOX2 pathway in colorectal cancer. J Exp Clin Cancer Res. 2020 Nov 5;39(1):232. doi: 10.1186/s13046-020-01690-z. PMID: 33153498; PMCID: PMC7643364.

30. Tait C, Patel AH, Chen A, Li Y, Minacapelli CD, Rustgi V. Early-Onset Colorectal Cancer: Prevalence, Risk Factors, and Clinical Features Among Commercially Insured Adults in the United States. Cureus. 2023 Nov 26;15(11):e49432. doi: 10.7759/cureus.49432. PMID: 38152812; PMCID: PMC10751861.

31. Ugai T, Haruki K, Harrison TA, Cao Y, Qu C, Chan AT, Campbell PT, Akimoto N, Berndt S, Brenner H, Buchanan DD, Chang-Claude J, Fujiyoshi K, Gallinger SJ, Gunter MJ, Hidaka A, Hoffmeister M, Hsu L, Jenkins MA, Milne RL, Moreno V, Newcomb PA, Nishihara R, Pai RK, Sakoda LC, Slattery ML, Sun W, Amitay EL, Alwers E, Thibodeau SN, Toland AE, Van Guelpen B, Woods MO, Zaidi SH, Potter JD, Giannakis M, Song M, Nowak JA, Phipps AI, Peters U, Ogino S. Molecular Characteristics of Early-Onset Colorectal Cancer According to Detailed Anatomical Locations: Comparison With Later-Onset Cases. Am J Gastroenterol. 2023 Apr 1;118(4):712–726. doi: 10.14309/ajg.0000000000002171. Epub 2022 Dec 30. PMID: 36707929; PMCID: PMC10065351.

32. Ullah F, Pillai AB, Omar N, Dima D, Harichand S. Early-Onset Colorectal Cancer: Current Insights. Cancers (Basel). 2023 Jun 15;15(12):3202. doi: 10.3390/cancers15123202. PMID: 37370811; PMCID: PMC10296149.

33. Waddell O, Mclauchlan J, McCombie A, Glyn T, Frizelle F. Quality of life in early-onset colorectal cancer patients: systematic review. BJS Open. 2023 May 5;7(3):zrad030. doi: 10.1093/bjsopen/zrad030. PMID: 37151082; PMCID: PMC10165061.

34. Wei J, Zheng X, Li W, Li X, Fu Z. Sestrin2 reduces cancer stemness via Wnt/β-catenin signaling in colorectal cancer. Cancer Cell Int. 2022 Feb 11;22(1):75. doi: 10.1186/s12935-022-02498-x. PMID: 35148781; PMCID: PMC8840770.

35. Wickström M, Dyberg C, Milosevic J, Einvik C, Calero R, Sveinbjörnsson B, Sandén E, Darabi A, Siesjö P, Kool M, Kogner P, Baryawno N, Johnsen JI. Wnt/β-catenin pathway regulates MGMT gene expression in cancer and inhibition of Wnt signalling prevents chemoresistance. Nat Commun. 2015 Nov 25;6:8904. doi: 10.1038/ncomms9904. PMID: 26603103; PMCID: PMC4674781.

36. Wilson JJ, Mick R, Wei SJ, Rustgi AK, Markowitz SD, Hampshire M, Metz JM. Clinical trial resources on the internet must be designed to reach underrepresented minorities. Cancer J. 2006 Nov-Dec;12(6):475–81. doi: 10.1097/00130404-20061100000007. PMID: 17207317.

37. Xu Y, Pasche B. TGF-beta signaling alterations and susceptibility to colorectal cancer. Hum Mol Genet. 2007 Apr 15;16 Spec No 1(SPEC). doi: 10.1093/hmg/ddl486. PMID: 17613544; PMCID: PMC2637552.

38. Yamada HY, Xu C, Jones KL, O’Neill PH, Venkateshwar M, Chiliveru S, Kim HG, Doescher M, Morris KT, Manne U, Rao CV. Molecular disparities in colorectal cancers of White Americans, Alabama African Americans, and Oklahoma American Indians. NPJ Precis Oncol. 2023 Aug 19;7(1):79. doi: 10.1038/s41698-023-00433-5. PMID: 37598287; PMCID: PMC10439889.

39. Zhang L, Ren CF, Yang Z, Gong LB, Wang C, Feng M, Guan WX. Forkhead Box S1 mediates epithelial-mesenchymal transition through the Wnt/β-catenin signaling pathway to regulate colorectal cancer progression. J Transl Med. 2022 Jul 21;20(1):327. doi: 10.1186/s12967-022-03525-1. PMID: 35864528; PMCID: PMC9306048.

40. Zhang Y, Wang X. Targeting the Wnt/β-catenin signaling pathway in cancer. J Hematol Oncol. 2020 Dec 4;13(1):165. doi: 10.1186/s13045-020-00990-3. PMID: 33276800; PMCID: PMC7716495.

41. Zhou W, Yan K, Xi Q. BMP signaling in cancer stemness and differentiation. Cell Regen. 2023 Dec 5;12(1):37. doi: 10.1186/s13619-023-00181-8. PMID: 38049682; PMCID: PMC10695912.

